# Unraveling the factors associated with digital health intervention uptake

**DOI:** 10.1101/2024.03.18.24304467

**Authors:** Ilona Ruotsalainen, Mikko Valtanen, Riikka Kärsämä, Adil Umer, Hilkka Liedes, Suvi Parikka, Annamari Lundqvist, Kirsikka Aittola, Suvi Manninen, Jussi Pihlajamäki, Anna-Leena Vuorinen, Jaana Lindström

**Affiliations:** VTT Technical Research Centre of Finland Ltd., Finland; Population Health Unit, Finnish Institute for Health and Welfare, Helsinki, Finland; Department of Mathematics and Statistics, University of Turku, Turku, Finland; Institute of Public Health and Clinical Nutrition, School of Medicine, University of Eastern Finland, Kuopio, Finland; Department of Medicine, Endocrinology and Clinical Nutrition, Kuopio University Hospital, Kuopio, Finland; Faculty of Social Sciences, Unit of Health Sciences, Tampere University, Tampere, Finland

## Abstract

For preventing chronic diseases, reducing modifiable lifestyle risk factors is crucial. Digital health interventions (DHIs) hold promise for reaching large population groups, but raise health equity concerns. A subgroup (n=6978) of 20 to 74-year-old participants of the population-based Healthy Finland survey were offered an opportunity to start using a web-based DHI, aiming to support adoption of healthy lifestyle habits. We used adjusted logistic regression models to identify significant predictors of DHI uptake. Women (adjusted odds ratio [aOR] 1.69, 95% CI 1.49–1.93), middle-aged individuals (1.47, 1.21–1.79), and those with higher income (aORs between 1.76–1.97) and education (1.10, 1.08–1.12) had higher odds of DHI uptake. Moreover, healthier lifestyle indicated by better diet quality (1.07, 1.04– 1.10), less frequent or non-smoking (aORs between 1.59–2.29), sleep (0.58, 0.37–0.86), higher functional capacity (1.06, 1.02–1.11), and good overall current health (1.46, 1.15– 1.89) associated with increased likelihood of DHI uptake. Lastly, individuals with better internet connections, higher competence to use e-services (aORs between 2.00–4.10), low concerns about data security (aORs between 1.37–1.76), stronger belief in benefits of digital services (1.04, 1.02–1.05), and reporting to use e-services (aORs between 2.48–6.08) had higher odds of uptake. Our findings indicate that those with higher socioeconomic status, better health and healthier lifestyle are more likely to take up DHI. Our research also highlights the impact of digital literacy and positive attitudes towards online services in adopting DHIs. These insights will be valuable in shaping strategies for future DHI initiatives.

## Introduction

Chronic noncommunicable diseases (NCDs) represent a significant global health challenge, imposing an increasing economic burden on healthcare services and resulting in premature deaths ^1,2^. To reduce the burden of NCDs, reducing modifiable lifestyle risk factors is highlighted as one of the most important actions globally ^3^. Adopting a healthy lifestyle can potentially reduce the risk of developing common chronic diseases by up to 80% ^4^. There is a pressing need to develop strategies that promote widespread adoption of healthy lifestyle habits, aiming to achieve impactful results at large scale.

Digital health interventions (DHIs) offer a viable way to support a healthy lifestyle and hold promise as a low-cost means to deliver interventions to large population groups. However, digital health interventions can be effective only if they reach the target population and those that need the intervention most. Ensuring that DHIs reach their intended users requires awareness of the potential barriers of DHI uptake. Currently, there is a lack of comprehensive understanding regarding the factors influencing DHI uptake. Identifying these factors will facilitate the customization of future interventions and recruitment strategies, thereby enhancing adoption rates and promoting digital equity.

Digital health promotion is an emerging field and therefore research on DHIs remains scarce ^5^. Previous research has mainly focused on the usage of health apps instead of DHIs, revealing that females ^6–8^, young or middle-aged individuals ^6–8^, as well as those with higher education levels ^6,7,9^, and higher income ^7,9,10^ tend to be more frequent users of health apps. Additionally, conflicting findings exist regarding the relation between overall health ^9,11,12^, lifestyle habits ^6,9,11,12^ and health app usage. Resent research has also suggested that additional factors like perceived usefulness and privacy concerns might play significant role in influencing the use of health apps and digital health services ^13–15^.

Studying health app users, while informative, does not offer comprehensive insights into potential barriers to DHI uptake. Firstly, the lack of comparative analysis between adopters and non-adopters raises questions about whether observed characteristics reflect a higher uptake probability or the characteristics of the populations to which the apps were advertised. Secondly, the usage of health apps itself may influence individuals’ lifestyle habits and health introducing a potential bias and thereby complicating the interpretation of these findings in relation to DHI adoption. Lastly, usage is also influenced by other factors such as user experience that may be affected by the app quality.

Here, our aim was to study factors predicting DHI uptake to identify potential groups that may less likely start using the DHIs. We examined whether socio-economic factors, self-reported health and lifestyle habits, or habitual use of e-services predict DHI uptake in a group that consisted of respondents to a survey offered for a representative random sample of Finnish population. Understanding these factors will help to identify population groups that are underrepresented in DHIs and find alternative approaches for them. These findings will also aid in integrating potential barriers into the formulation of recruitment strategies and the development of DHIs themselves.

## Results

### Baseline information

Of the final sample of 6975 participants, 1287 (18.4%) started using the app (“adopters”). Among those who were invited with SMS the uptake proportion was 16.2% (n = 806/4975) and among those with letter invitation 24.1% (n = 481/2000). Those invited via SMS were less likely to start using the app than those invited via mail (aOR 0.65 [0.55–0.75]). Background information for the whole study sample and separately for adopters and non-adopters is presented in Table 1.

**Table 1.** Background information for the study sample.

|  | **All** |  | **Adopters** |  | **Non-adopters** |  |
| --- | --- | --- | --- | --- | --- | --- |
| | Mean ± SD / N (%) | Missing | Mean ± SD / N (%) | Missing | Mean ± SD / N (%) | Missing |
| ***Socio-economics*** |  |  |  |  |  |  |
| **Age (years)** |  | 0 |  | 0 |  | 0 |
| 20 – 34 | 1229 (18%) |  | 250 (19%) |  | 979 (17%) |  |
| 35 – 49 | 1254 (18%) |  | 303 (24%) |  | 951 (17%) |  |
| 50 – 64 | 2066 (30%) |  | 361 (28%) |  | 1705 (30%) |  |
| 65 – 74 | 2426 (35%) |  | 373 (29%) |  | 2053 (36%) |  |
| **Sex** |  | 0 |  | 0 |  | 0 |
| Female | 3975 (57%) |  | 868 (67%) |  | 3107 (55%) |  |
| Male | 3000 (43%) |  | 419 (33%) |  | 2581 (45%) |  |
| **Education (years)** | 14.0 ± 3.6 | 95 | 15.2 ± 3.4 | 10 | 13.8 ± 3.6 | 85 |
| **Household income (euros per year)** |  | 95 |  | 11 |  | 84 |
| < 15 000 € | 761 (11%) |  | 94 (7%) |  | 667 (12%) |  |
| 15 001 – 35 000 € | 1913 (28%) |  | 266 (21%) |  | 1647 (29%) |  |
| 35 001 – 55 000 € | 1916 (28%) |  | 380 (30%) |  | 1536 (27%) |  |
| 55 001 – 75 000 € | 1171 (17%) |  | 264 (21%) |  | 907 (16%) |  |
| > 75 000 € | 1119 (16%) |  | 272 (21%) |  | 847 (15%) |  |
| **Employment status** |  | 48 |  | 2 |  | 46 |
| Employed full-time | 1798 (42%) |  | 389 (48%) |  | 1409 (41%) |  |
| Employed part-time | 211 (5%) |  | 41 (5%) |  | 170 (5%) |  |
| Retired | 1463 (35%) |  | 228 (28%) |  | 1235 (36%) |  |
| On a disability pension or rehabilitation benefit | 161 (4%) |  | 24 (3%) |  | 137 (4%) |  |
| Part-time retirement | 27 (1%) |  | 1 (0%) |  | 26 (1%) |  |
| Unemployed | 169 (4%) |  | 36 (4%) |  | 133 (4%) |  |
| Family leave or stay-at-home parent | 67 (2%) |  | 19 (2%) |  | 48 (1%) |  |
| Other | 123 (3%) |  | 21 (3%) |  | 102 (3%) |  |
| **Health** |  |  |  |  |  |  |
| **BMI (kg/m<sup>2</sup>)** | 27.4 ± 5.3 | 75 | 27.5 ± 5.5 | 6 | 27.3 ± 5.2 | 69 |
| **Functional capacity** | 13.8 ± 2.0 | 144 | 14.1 ± 1.5 | 15 | 13.7 ± 2.1 | 129 |
| **Ability to work** |  | 59 |  | 5 |  | 54 |
| Completely unable to work | 350 (5%) |  | 49 (4%) |  | 301 (5%) |  |
| Partially unable to work | 1688 (24%) |  | 247 (19%) |  | 1441 (26%) |  |
| Completely fit for work | 4878 (71%) |  | 986 (77%) |  | 3892 (69%) |  |
| **Current health status** |  | 39 |  | 1 |  | 38 |
| Poor or fairly poor | 676 (10%) |  | 83 (6%) |  | 593 (11%) |  |
| Average | 1740 (25%) |  | 278 (22%) |  | 1462 (26%) |  |
| Good or fairly good | 4520 (65%) |  | 925 (72%) |  | 3595 (64%) |  |
| **Long term illnesses** |  | 106 |  | 12 |  | 94 |
| Yes | 3937 (57%) |  | 713 (56%) |  | 3224 (58%) |  |
| No | 2932 (43%) |  | 562 (44%) |  | 2370 (42%) |  |
| **Health care visits** | 5.7 ± 7.4 | 655 | 5.8 ± 6.6 | 125 | 5.6 ± 7.6 | 530 |
| **Ability to learn new** |  | 55 |  | 3 |  | 52 |
| Poorly or very poorly | 277 (4%) |  | 34 (3%) |  | 243 (4%) |  |
| Adequately | 1779 (26%) |  | 249 (19%) |  | 1530 (27%) |  |
| Well or very well | 4864 (70%) |  | 1001 (78%) |  | 3863 (69%) |  |
| **Limitations due to health problems** |  | 135 |  | 20 |  | 115 |
| Severely limited | 384 (6%) |  | 51 (4%) |  | 333 (6%) |  |
| Limited but not severely | 2424 (35%) |  | 413 (33%) |  | 2011 (36%) |  |
| Not limited at all | 4032 (59%) |  | 803 (63%) |  | 3229 (58%) |  |
| **Lifestyle habits** |  |  |  |  |  |  |
| **Diet quality** | 8.1 ± 2.5 | 193 | 8.5 ± 2.5 | 31 | 7.9 ± 2.5 | 162 |
| **Physical activity** |  | 323 |  | 31 |  | 292 |
| Did not achieve recommended level | 4122 (62%) |  | 741 (59%) |  | 3381 (63%) |  |
| Achieved recommended level | 2530 (38%) |  | 515 (41%) |  | 2015 (37%) |  |
| **Enough sleep** |  | 50 |  | 6 |  | 44 |
| Yes, almost always or often | 5324 (77%) |  | 991 (77%) |  | 4333 (77%) |  |
| Rarely or hardly ever | 1315 (19%) |  | 261 (20%) |  | 1054 (19%) |  |
| Not sure | 286 (4%) |  | 29 (2%) |  | 257 (5%) |  |
| **Smoking** |  | 196 |  | 33 |  | 163 |
| Daily | 675 (10%) |  | 56 (4%) |  | 619 (11%) |  |
| Occasionally | 419 (6%) |  | 64 (5%) |  | 355 (6%) |  |
| Not at all | 3137 (46%) |  | 586 (47%) |  | 2551 (46%) |  |
| Have never smoked | 2548 (38%) |  | 548 (44%) |  | 2000 (36%) |  |
| **Use of e-services** |  |  |  |  |  |  |
| **Independence of e-service use** |  | 45 |  | 4 |  | 41 |
| Do not use | 447 (6%) |  | 11 (1%) |  | 436 (8%) |  |
| Use with help or someone else uses on my behalf | 306 (4%) |  | 20 (2%) |  | 286 (5%) |  |
| Use independently | 6177 (89%) |  | 1252 (98%) |  | 4925 (87%) |  |
| **Competence to use online services** |  | 62 |  | 1 |  | 61 |
| No competence | 208 (3%) |  | 6 (0%) |  | 202 (4%) |  |
| Low competence | 550 (8%) |  | 37 (3%) |  | 513 (9%) |  |
| Moderate competence | 2076 (30%) |  | 268 (21%) |  | 1808 (32%) |  |
| High competence | 1976 (29%) |  | 446 (35%) |  | 1530 (27%) |  |
| Very high competence | 2103 (30%) |  | 529 (41%) |  | 1574 (28%) |  |
| **Non-accessibility of electronic services** |  | 539 |  | 57 |  | 482 |
| Completely or somewhat agree | 831 (13%) |  | 118 (10%) |  | 713 (14%) |  |
| Neither agree nor disagree | 1583 (25%) |  | 274 (22%) |  | 1309 (25%) |  |
| Strongly or somewhat disagree | 4022 (63%) |  | 838 (68%) |  | 3184 (61%) |  |
| **Concerned about data security** |  | 294 |  | 24 |  | 270 |
| Completely agree | 724 (11%) |  | 80 (6%) |  | 644 (12%) |  |
| Somewhat agree | 1441 (22%) |  | 255 (20%) |  | 1186 (22%) |  |
| Neither agree nor disagree | 1242 (19%) |  | 203 (16%) |  | 1039 (19%) |  |
| Somewhat disagree | 1392 (21%) |  | 310 (25%) |  | 1082 (20%) |  |
| Strongly disagree | 1882 (28%) |  | 415 (33%) |  | 1467 (27%) |  |
| **Poor internet connections in the area** |  | 378 |  | 29 |  | 349 |
| Completely or somewhat agree | 727 (11%) |  | 111 (9%) |  | 616 (12%) |  |
| Neither agree nor disagree | 1050 (16%) |  | 151 (12%) |  | 899 (17%) |  |
| Strongly or somewhat disagree | 4820 (73%) |  | 996 (79%) |  | 3824 (72%) |  |
| **Benefits of digital services** | 16.4 ± 5.1 | 465 | 17.4 ± 4.7 | 54 | 16.1 ± 5.2 | 411 |

For continuous variables, data are mean ± SD and for categorical variables data are N (%). BMI= body mass index, SD = standard deviation.

### Predictor variables associated with the uptake of the DHI

Factors associated with uptake of BitHabit application were studied in five domains: socio-economics, health, lifestyle habits, and use of e-services. Adjusted odds ratios (aOR) from all the models along with associated p-values and 95% confidence intervals are listed in Table 2 and plotted in Fig 1.

**Table 2.** Adjusted ORs for predictor variables.

| Variable | LRT |  | Adjusted OR |  |
| --- | --- | --- | --- | --- |
| | $\chi^2$ | FDR adjusted P-value | aOR (95% CI) | P-value (group comparison) |
| ***Socio-economics*** |  |  |  |  |
| **Age (years)** | 22.37 | 0.0001 |  |  |
| 20 – 34 (ref) |  |  |  |  |
| 35 – 49 |  |  | 1.47 (1.21–1.79) | 0.0001 |
| 50 – 64 |  |  | 1.12 (0.91–1.37) | 0.282 |
| 65 – 75 |  |  | 1.00 (0.81–1.22) | 0.993 |
| **Sex** | 66.75 | <0.0001 |  |  |
| Male (ref) |  |  |  |  |
| Female |  |  | 1.69 (1.49–1.93) | <0.0001 |
| **Education (years)** | 98.80 | <0.0001 | 1.10 (1.08–1.12) |  |
| **Household income (euros per year)** | 50.19 | <0.0001 |  |  |
| < 15 000 € (ref) |  |  |  |  |
| 15 001 – 35 000 € |  |  | 1.20 (0.93–1.57) | 0.164 |
| 35 001 – 55 000 € |  |  | 1.76 (1.37–2.27) | <0.0001 |
| 55 001 – 75 000 € |  |  | 1.97 (1.51–2.59) | <0.0001 |
| > 75 000 € |  |  | 1.96 (1.50–2.59) | <0.0001 |
| **Employment status** | 3.60 | 0.891 |  |  |
| Employed full-time (ref) |  |  |  |  |
| Employed part-time |  |  | 0.97 (0.73–1.29) | 0.846 |
| Retired |  |  | 0.93 (0.64–1.34) | 0.713 |
| On a disability pension or rehabilitation benefit |  |  | 0.95 (0.67–1.34) | 0.793 |
| Part-time retirement |  |  | 0.77 (0.31–1.63) | 0.522 |
| Unemployed |  |  | 1.11 (0.79–1.53) | 0.526 |
| Family leave or stay-at-home parent |  |  | 0.78 (0.47–1.24) | 0.316 |
| Other |  |  | 1.00 (0.68–1.45) | 0.992 |
| Student |  |  | 1.18 (0.87–1.58) | 0.281 |
| **Health** |  |  |  |  |
| **BMI (kg/m<sup>2</sup>)** | 3.60 | 0.076 | 1.01 (1.00–1.02) |  |
| **Functional capacity** | 9.27 | 0.005 | 1.06 (1.02–1.11) |  |
| **Ability to work** | 1.16 | 0.612 |  |  |
| Completely unable to work (ref) |  |  |  |  |
| Partially unable to work |  |  | 1.03 (0.75–1.44) | 0.861 |
| Completely fit for work |  |  | 0.94 (0.67–1.33) | 0.716 |
| **Current health status** | 11.99 | 0.005 |  |  |
| Poor or fairly poor (ref) |  |  |  |  |
| Average |  |  | 1.24 (0.95–1.63) | 0.119 |
| Good or fairly good |  |  | 1.46 (1.15–1.89) | 0.003 |
| **Long term illnesses** | 0.25 | 0.644 |  |  |
| Yes (ref) |  |  |  |  |
| No |  |  | 0.97 (0.85–1.10) | 0.616 |
| **Health care visits** | 0.41 | 0.602 | 1.00 (0.99–1.01) |  |
| **Ability to learn new** | 6.24 | 0.063 |  |  |
| Poorly or very poorly (ref) |  |  |  |  |
| Adequately |  |  | 0.99 (0.68–1.49) | 0.957 |
| Well or very well |  |  | 1.20 (0.84–1.78) | 0.334 |
| **Limitations due to health problems** | 3.06 | 0.263 |  |  |
| Severely limited (ref) |  |  |  |  |
| Limited but not severely |  |  | 1.07 (0.79–1.50) | 0.661 |
| Not limited at all |  |  | 1.19 (0.88–1.65) | 0.268 |
| **Lifestyle** |  |  |  |  |
| **Diet quality** | 23.67 | <0.0001 | 1.07 (1.04–1.10) |  |
| **Physical activity** | 3.34 | 0.086 |  |  |
| Did not achieve recommended level (ref) |  |  |  |  |
| Achieved recommended level |  |  | 1.13 (0.99–1.29) | 0.067 |
| **Enough sleep** | 7.54 | 0.038 |  |  |
| Rarely or hardly ever (ref) |  |  |  |  |
| Yes, almost always or often |  |  | 0.96 (0.82–1.13) | 0.623 |
| Not sure |  |  | 0.58 (0.37–0.86) | 0.010 |
| **Smoking** | 41.76 | <0.0001 |  |  |
| Daily (ref) |  |  |  |  |
| Occasionally |  |  | 1.59 (1.08–2.36) | 0.020 |
| Not at all |  |  | 2.26 (1.69–3.06) | <0.0001 |
| Have never smoked |  |  | 2.29 (1.72–3.12) | <0.0001 |
| **Use of e-services** |  |  |  |  |
| **Independence of e-service use** | 70.22 | <0.0001 |  |  |
| Don't use (ref) |  |  |  |  |
| Use with help or someone else uses on my behalf |  |  | 2.48 (1.19–5.47) | 0.018 |
| Use independently |  |  | 6.08 (3.46–11.92) | <0.0001 |
| **Competence to use online services** | 97.61 | <0.0001 |  |  |
| No or low competence (ref) |  |  |  |  |
| Moderate competence |  |  | 2.00 (1.44–2.85) | <0.0001 |
| High competence |  |  | 3.49 (2.50–4.98) | <0.0001 |
| Very high competence |  |  | 4.10 (2.90–5.92) | <0.0001 |
| **Non-accessibility of electronic services** | 6.41 | 0.062 |  |  |
| Completely or somewhat agree (ref) |  |  |  |  |
| Neither agree nor disagree |  |  | 1.03 (0.81–1.31) | 0.831 |
| Strongly or somewhat disagree |  |  | 1.22 (0.98–1.52) | 0.082 |
| Concerned about data security | 24.75 | 0.0001 |  |  |
| Completely agree (ref) |  |  |  |  |
| Somewhat agree |  |  | 1.46 (1.11–1.93) | 0.007 |
| Neither agree nor disagree |  |  | 1.37 (1.03–1.82) | 0.030 |
| Somewhat disagree |  |  | 1.76 (1.35–2.33) | <0.0001 |
| Strongly disagree |  |  | 1.76 (1.35–2.31) | <0.0001 |
| **Poor internet connections in the area** | 11.11 | 0.007 |  |  |
| Completely or somewhat agree (ref) |  |  |  |  |
| Neither agree nor disagree |  |  | 0.91 (0.70–1.20) | 0.509 |
| Strongly or somewhat disagree |  |  | 1.22 (0.98–1.53) | 0.074 |
| **Benefits of digital services** | 27.38 | <0.0001 | 1.04 (1.02–1.05) |  |

BMI = body mass index, FDR = false discovery rate, LRT = likelihood ratio test, aOR = adjusted odds ratio. Details of the adjustments for each variable are listed in methods section.

**Fig. 1:**
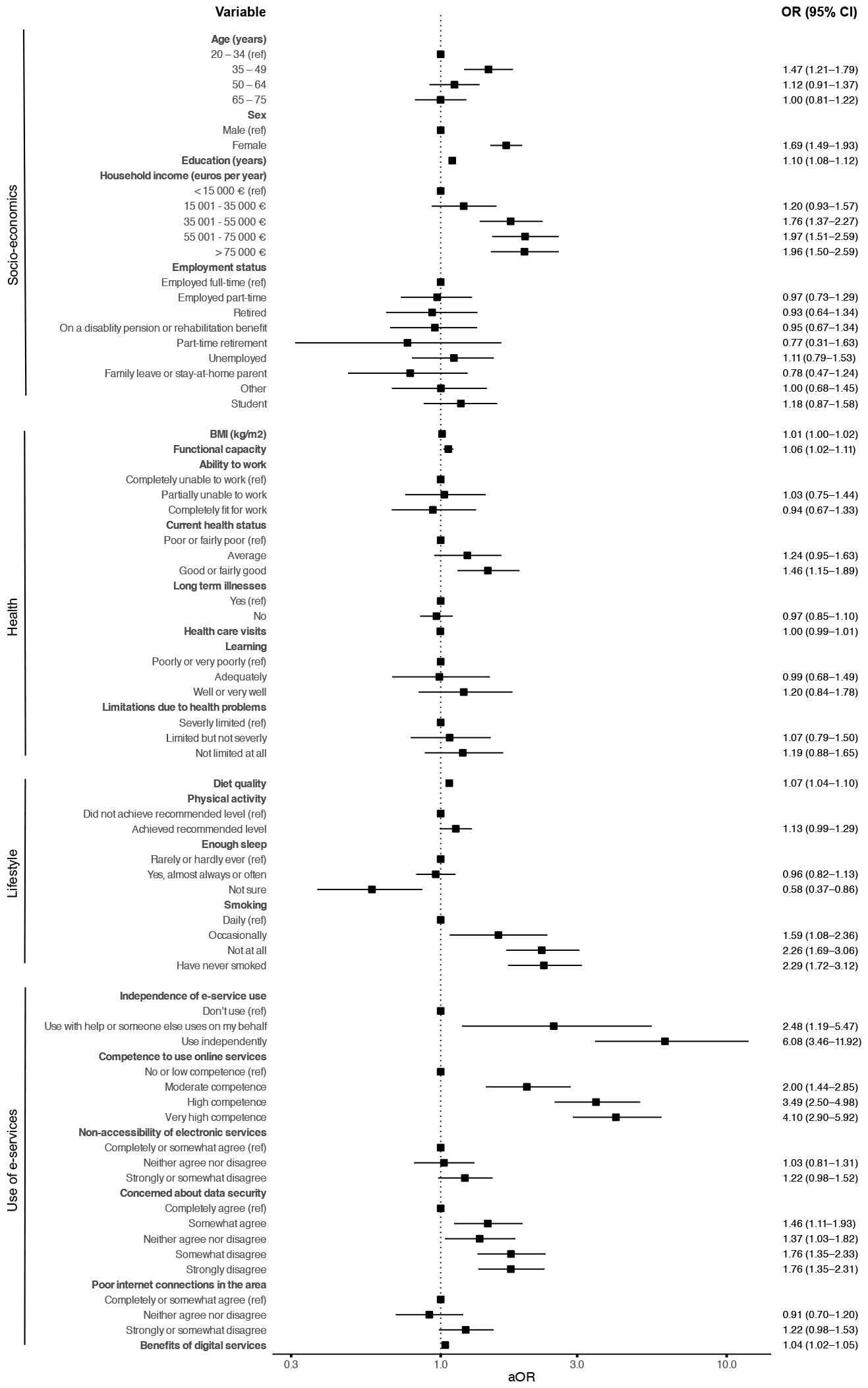
Forest plots for adjusted ORs with 95% CI for predictor variables related to the uptake of the BitHabit app. aOR = adjusted odds ratio, BMI = body mass index, CI = confidence interval. Details of the adjustments for each variable are listed in statistical methods section.

#### Socio-economics

Age, sex, education, and income demonstrated a significant association with the uptake of the DHI (Table 2). Odds of adopting the DHI for age group 35–49 years were higher compared to the reference group of 20–34 years (aOR 1.47, [95% CI 1.21–1.79]). The differences of the two older age groups against the reference group were small and themselves not significant. Longer education in years was related to greater odds for adopting the DHI (aOR 1.10 [95% CI 1.08–1.12]). In the income groups, higher income was associated with greater odds of adopting the DHI compared to the reference group (35 000€ or less), though the estimates for the two highest categories were virtually identical (35 001– 55 000€: aOR 1.76 [1.37–2.27]; 55 001–75 000€: aOR 1.97 [1.51–2.59]; >75 000€: aOR 1.96 [1.50–2.59]). In addition, women were more likely than men to adopt the DHI (aOR 1.69 [1.49–1.93]). Employment status was not significantly associated with DHI uptake.

#### Health

Of the health-related predictors, functional capacity and current health status showed a significant association with the DHI uptake (Table 2). The odds of adopting the DHI increased with higher functional status (aOR 1.06 [95% CI 1.02–1.11]) and better current health (good or fairly good compared to poor: aOR 1.46 [1.15–1.89]). BMI, ability to work, ability to learn new, presence of long-term illnesses, the number of healthcare visits during past 12 months, and limitations due to health problems were not significantly associated with uptake of the DHI.

#### Lifestyle habits

Diet quality, smoking, sleeping, and lifestyle were associated with the uptake of the DHI (Table 2). Higher odds to DHI uptake were estimated for better diet quality (aOR 1.07 [1.04– 1.10]) and less frequent smoking (occasionally vs. daily: aOR 1.59 [1.08–2.36]; not at all vs. daily: aOR 2.26 [1.69–3.06]; having never smoked vs. daily: aOR 2.29 [1.72–3.12]). Sleeping enough was also a significant predictor, however, no meaningful difference was estimated between those sleeping mostly enough and the reference category of those sleeping mostly not enough. Instead, lower odds were obtained for the group being unsure whether they sleep enough (compared to reference category: aOR 0.58 [0.37–0.86]). Achieving physical activity recommendations was not significantly associated with uptake of the DHI.

#### Use of e-services

Each predictor in the use of e-services domain (competence to use online services, independence of e-service use, data security concerns, quality of data connections, and belief in the benefits of digital services) except the perceived accessibility of electronic services showed a significant connection with the DHI uptake (Table 2). Higher odds for adoption of BitHabit DHI were found in those using e-services (use with help or someone else uses on my behalf vs. do not use: aOR, 2.48 [1.19–5.47]; use independently vs. do not use: aOR 6.08 [3.46–11.92]), greater competence of using online services (moderate vs. no or low competence: aOR 2.00 [1.44–2.85]; high vs. no or low competence: aOR 3.49 [2.50– 4.98]; very high vs. no or low competence: aOR 4.10 [2.90–5.92]), reduced concerns about data security (somewhat agree that I am concerned about data security vs. completely agree: aOR1.46 [1.11–1.93]; neither agree nor disagree vs. completely agree: aOR 1.37 [1.03–1.82]; somewhat disagree vs. completely agree: aOR 1.76 [1.35–2.33]; strongly disagree vs. completely agree 1.76 [1.35–2.31]), and stronger belief in the benefits of digital services (aOR 1.04 [1.02–1.05]). Regarding quality of internet connections, the individuals indicating good connections were estimated to have the greatest odds of adopting the DHI.

## Discussion

This study examined relation of background characteristics in four different domains (socio-economics, health, lifestyle habits, and use of e-services) with uptake of lifestyle DHI. Overall, 18.4% of the participants who received the invitation started using the BitHabit application. We found that women, middle-aged individuals, and those with higher income and education were more likely to take up the DHI. Moreover, better lifestyle choices as well as better functional capacity and current health were positively associated with the higher odds of taking up the DHI. Our research also underscores the impact of digital literacy and positive attitudes towards online services, revealing that individuals with greater familiarity and more positive views about online services, were more likely to register to use the DHI. These findings imply that when offering DHIs, it may be challenging to engage those individuals that would benefit the most from lifestyle DHI, emphasizing the importance of tailoring recruitment and intervention methods accordingly.

While there is a scarcity in studies directly comparing DHI adopters and non-adopters, several studies exploring health app usage report that women and individuals in the young or middle-aged categories use more health apps ^6,7,16,17^. These findings are also supported by recent results from a large population-level study, comparing the characteristics of individuals who registered to use a mobile health physical activity intervention (n = 696 907) with the entire population of Singapore, showing that female and younger participants were more inclined to register for health app ^8^. Our findings that women and those aged 35–49 years are more likely to take up the DHI are mainly consistent with these previous studies examining health app usage. Interestingly, our results do not support the idea that young adults are more likely to take up DHI. These conflicting findings could be due to differences in study design as most of the previous studies examining usage focus on consumer-targeted health applications which may be more appealing to younger adults. Regarding income and education, previous studies have proposed that individuals with higher income levels or socioeconomic status and those with higher levels of education tend to utilize more health apps ^6,7,9,10^. The current results showing that those with higher income levels and higher education are more likely to take up the DHI align with the previous findings on health app usage.

DHIs and digital lifestyle interventions aim to promote healthy lifestyle and improve health outcomes. However, in general, it is not well known whether individual’s lifestyle or health influences uptake of DHI. Our results propose that better diet quality and smoking less or not at all as well as better overall health, as indicated by self-reported functional capacity and current health status are related to higher odds of taking up DHI. In this study perceived lack of sleep was not associated with uptake of the DHI; however, those individuals who were not sure whether they get enough sleep had lower odds for taking up the DHI. This response might indicate a lack of personal interest in the individual’s lifestyle habits, however, it is challenging to draw strong conclusions based on this answer. Prior studies imply a potential link between health app usage and lifestyle choices, but results regarding diet quality, smoking, and physical activity still remain inconclusive in earlier investigations ^6,9,11,12,18^. Conflicting results have also been found on the link between health app use and BMI ^8,9,12^ as well as between chronic diseases and health app possession ^9,11,12,19^. These earlier contradictory findings may be due to variations in the study design. Interestingly, those with better self-reported health status have been reported to be more likely to download health apps ^19^. Overall, the results of the current study regarding lifestyle habits and health imply that if DHI aims to target those with poorer health and non-healthy lifestyle, we may not get the intended target population involved when employing a one-size-fits-all recruitment strategy.

The current findings also indicate that digital competence and views about online service impact the uptake of DHI. In particular, individuals’ competence and their ability to use e-services independently are closely linked to their likelihood of taking up DHI. Those reporting very high competence in utilizing e-services had a four-fold odds to register to DHI compared to those with little to no competence. Additionally, individuals who independently used e-services had a six-fold odds to take up the DHI compared to those who did not use e-services at all. These results highlight the importance of addressing the needs of individuals with limited e-service competence to enable them to access DHI, or alternatively, exploring alternative approaches. Furthermore, concerns about data security impacted the odds of DHI uptake. Previous research on digital health services ^13,20^ and mobile health apps ^14,15^ has also highlighted the role of data security concerns as potential barriers to the usage of these services. Addressing security-related issues, such as ensuring compliance with regulations and enhancing transparency regarding data collection and handling, may help alleviate some of these concerns.

DHIs have significant potential to deliver interventions to large population groups and therefore possibility to decrease the burden of NCDs on population level. Despite the potential, there are notable concerns regarding the digital divide and its impact on health equity ^21^. Our findings indicate those with higher socioeconomical status, greater digital literacy, and better health and lifestyle habits were more likely to take up DHI. Therefore, effectively reaching individuals likely to benefit most from lifestyle DHIs may be complex. Consequently, it is important to design the recruitment strategies and the intervention modalities considering the targeted population group. Otherwise the digitalization of lifestyle interventions might lead to widening health equity gaps between population groups.

Our approach has some strengths and limitations. Most importantly, our study population was drawn among participants of a recent population based survey covering wide age and socio-economic distribution, and the available knowledge of the background characteristics of the whole approached population enabled a direct comparison between adopters and non-adopters, instead of only studying adopters that is a common approach in earlier research. There are also some potential limitations. It is worth noting that individuals who had chosen not to participate in the Healthy Finland survey were excluded from the sample used in the current study. Also, the Healthy Finland survey was firstly asked to be filled online, which could introduce bias in the respondents who were later on approached and offered the DHI. Nevertheless, this bias may have been mitigated by the option to request a paper format for the survey. This study was conducted only in one country (Finland), which might limit the generalizability of the results in other countries. Specifically, the significance of the quality of internet connections in the area may be more pronounced in regions with a higher proportion of individuals experiencing poor connections.

## Methods

### Participants and study design

This cross-sectional study was a sub-study of a Healthy Finland population survey ^22^. In this survey a questionnaire on health, well-being and service use was sent to randomly selected persons over the age of 20, representing the entire adult population of Finland. Finnish speaking individuals aged 20 to 74 years who had answered the Healthy Finland questionnaire between September 2022 and the end of 2022 and who were not invited to participate to a health examination part of the Healthy Finland survey, were considered eligible for this current study. An SMS invitation was sent to all eligible individuals with a known phone number (n=4978 [71%]). Additionally, 2000 (29%) individuals from the rest of the eligible population were sampled based on 5-year age-groups to receive a letter invitation. Three participants asked their data to be removed resulting in a final sample size of 6975 individuals. The sample size was chosen to detect small effects in logistic regression with statistical power of 0.8, type I error of 0.05, and anticipated that approximately 10–15% of the invitees would adopt the DHI.

The individuals were offered an opportunity to use a DHI application (BitHabit app) for three months ^23^. The invitation letter included web address and SMS message a direct link to the project’s web page (hosted by Finnish Institute for Health and Welfare) where information regarding the study, a link to the BitHabit app, and brief instructions on how to begin the use of the BitHabit app were given. The participation to the study was voluntary and based on informed consent. The ethical approval for the study was obtained from the research ethics committee of the Finnish Institute for Health and Welfare (THL/5335/6.02.01/2022).

### Digital health intervention app (BitHabit)

The BitHabit app was originally developed to provide support for formation of healthy lifestyle habits in adults at an increased risk of type 2 diabetes ^23,24^. It includes a broad selection of small health promoting actions (“habits”) in 14 different categories related to physical activity, diet, sleep, stress management, positive mood, smoking, and alcohol consumption. The BitHabit app is a web-based application that can be used with a mobile phone, tablet or computer. To log into the app, the users had to provide a phone number and a user id that was given either in the letter or SMS message. The uptake of the app was defined as accepting the invitation to participate, agreeing to the terms of the BitHabit app, and registration to the app with a phone number and user id.

### Predictor variables

Predictors from the following categories were examined in the current study: socio-economics, health, lifestyle habits, and use of electronic services. All predictor variables except age and sex (assigned at birth), which were obtained from Finnish National Population Register, were obtained from the Healthy Finland survey questionnaire and were thus self-reported. The survey was conducted approximately two to four months prior to sending invitations to take part to the DHI sub-study. All questions and the answer options are presented in Supplementary material (p 1).

#### Socio-economics

Socio-economic variables included age, sex, education (years of education including primary and comprehensive school), household income (euros per year before taxes) and employment status. The household income was coded as a 5-category ordinal variable (< 15 000 €, 15 001 – 35 000 €, 35 001 – 55 000 €, 55 001 – 75 000 € and > 75 000 €). Age was categorized into four classes (20 – 34, 35 – 49, 50 – 64, 65 – 74).

#### Health

Body mass index (BMI) was calculated based on self-reported height and weight. Other health-related predictor variables included functional capacity, ability to work, current health status, prevalence of (any) long term illness, health care service use (number of visits during past 12 months), perceived ability to learn and perceived limitations due to health problems. Functional capacity was calculated as a summary score for five questions on the abilities to perform the following activities: run about 100 m; walk about 500 m without stopping to rest; see ordinary newspaper print; hear what is said in a conversation between several people; and walk up one flight of stairs without stopping to rest. Each sub question had 4 choices (yes, with no problem [3 points] / yes, with some difficulty [2 points] / yes, but with great difficulty [1 point] / no, I cannot [0 points]). A higher summary score indicated better functional capacity.

#### Lifestyle habits

Lifestyle-related information included physical activity, sleep, smoking and diet quality. The participants were divided into two groups based on whether they achieved the Finnish physical activity recommendations ^25^. Sleep adequacy categories were created based on the question “do you get enough sleep”. Diet quality score was calculated following the method presented in Lindström et al. ^26^ and the details of the scoring are presented in Supplementary material (p 4).

#### Use of electronic services

Following predictors relating to the use of e-services were considered: independence of e-service use, competence to use e-services, accessibility of e-services, concerns about data security, poor internet connections and perceived benefits of e-services. Perceived benefits of e-services score refers to a summary score calculated based on the level of agreement to six claims: e-services 1) help me to assess the need for services, 2) support me in finding and choosing the most suitable services, 3) make it easier for me to use services regardless of where I am and when, 4) make it easier for me to collaborate with professionals, 5) help me to take an active role in looking after my own health and welfare, and 6) help me to take care of the health, welfare and functional capacity of family or friends. Each claim had five categories (completely agree / somewhat agree / neither agree nor disagree / somewhat disagree / strongly disagree). A higher score indicated that a person experienced e-services more beneficial.

### Statistical methods

Logistic regression models were used to assess the association between the predictor variables and the uptake of the BitHabit DHI. The results are presented as adjusted odds ratios (aORs) with 95% confidence intervals (CIs). Models examining the variables related to health, lifestyle, and use of e-services were adjusted for contact method (SMS or letter), age, sex, education, and income. Regarding socio-economic factors, models for age, sex, and contact method were adjusted by each other; models for income, BMI and employment status by education, contact method, age, and sex; the model for education by contact method, age, and sex. Participants with missing data (missing answers) were excluded from the analyses and a complete-case analysis was performed. The amount of missing data for each predictor variable is presented in Results section Table 1.

The overall significance of the associations between the categorical predictors and the outcome were assessed with likelihood ratio tests (LRT), for which a false discovery rate (FDR) corrected P-value less than 0.05 was determined to indicate a significant association. The R software, version 4.3.1. ^27^ was used to perform all statistical analyses.

## Data Availability

In accordance with ethical approval of the study, individual participant data are not publicly available due to identifying and sensitive information.

## Contributions

JL, A-LV and IR designed the study. IR, AU, AL, SP were involved in the data collection and AU maintained the BitHabit software. Analysis plan was designed by MV, JL and IR and analyses were done by MV. IR, MV and JL interpreted the data and IR visualized the results. Original draft was written by IR, MV, RK and JL. All authors reviewed and edited the manuscript. JL and JP acquired the funding for this project.

## Acknowledgements

This study was funded by the Academy of Finland (grant numbers 332464, 332465, and 332466).

