## Supplementary material for "Unraveling the factors associated with digital health intervention uptake"

### Questions on predictor variables in Healthy Finland survey

Socio-economics

1. Education
   How many years altogether have you attended school or studied full time (in years)? Including primary and comprehensive school.

2. Income
   How large was your household's income last year (before taxes)?
   less than 15 000 € (less than 1250 €/month)
   15 001 - 35 000 € (about 1251–2915 €/month)
   35 001 - 55 000 € (about 2916–4580 €/month)
   55 001 - 75 000 € (about 4581–6250 €/month)
   more than 75 000 € (6251 €/month or more)

3. Employment situation
   At the moment, are you principally:
   Employed full-time
   Employed part-time _______ hours per week
   Retired on an old age pension
   Receiving a disability pension or rehabilitation benefit
   On part-time retirement
   Unemployed or laid off, length of current period in months: _______ months
   On family leave, or a stay-at-home mother/father
   A student or on study leave
   Other

Health

4. Ability to work
   Regardless of whether you are employed or not, please estimate your current work capacity. Are you:
   Completely fit for work
   Partially unable to work
   Completely unable to work?

5. Current health status
   How would you describe your state of health at present?
   Good or fairly good
   Average
   Fairly poor or poor

6. Prevalence of (any) long term illness
   Do you have any longstanding illness or longstanding health problem?
   Yes
   No

7. <u>Health care services use</u>
   Have you used health care services (e.g. doctor, nurse, hospital, dentist, dental hygienist) in the past 12 months?
   No
   Yes

8. <u>Perceived ability to learn</u>
   How easily do you learn new things?
   Very well or well
   Adequately
   Poorly or very poorly

9. <u>Perceived limitations due to health problems</u>
   Are you limited because of a health problem in activities people usually do?
   Severely limited
   Limited but not severely
   Not limited at all

Lifestyle

10. <u>Diet</u>
    How often do you usually consume the following foods and drinks?
    Fruit, berries (no juices)
    Vegetables, root vegetables (no potatoes)
    Legumes, plant protein products (e.g., peas, tofu, faba bean product)
    Whole grain bread/porridge, whole grain garnish (e.g., brown rice)
    Milk and milk products (e.g. cheese, yoghurt)
    Plant-based products used as an alternative to milk products (e.g., oat drink)
    Fish
    White meat (e.g. chicken)
    Red meat and meat products (e.g. cold cuts)
    Choices: not at all, 1-2 times a day, 2-6 times a week, once a week or less often, 3 times a day or more often

11. <u>Physical activity</u>
    How much exercise do you get in a week in the course of your work, commute, and spare time? Think about the past 12 months. Take all regular, weekly physical exertion in consideration.
    hardly any regular exercis.
    Low-intensity aerobic exercise (= does not make you sweat or get out of breath, e.g. walking leisurely) in total _______ hours and _______ minutes a week
    Moderate-intensity aerobic exercise (= makes you sweat a bit and/or get slightly out of breath, e.g. walking briskly) in total _______ hours and _______ minutes a week
    High-intensity aerobic exercise (= makes you sweat a lot and/or get out of breath, e.g. jogging or running) in total _______ hours and _______ minutes a week

12. <u>Sleep</u>
    Do you feel that you get enough sleep?

yes, almost always or yes, often
rarely or hardly ever
not sure

13. Smoking
    Do you smoke currently (cigarettes, cigars or pipe)?
    yes, daily
    occasionally
    not at all
    I have never smoked

Use of e-services

14. Use of e-services
    Do you use the Internet to access e-services (e.g. My Kanta, MyTax, OmaKela)?
    I use it independently
    I use it with another person's help or someone else uses it on my behalf
    I don't use it

15. Competence to use e-services
    How would you rate your competence to use online services (on a computer or smartphone)?
    No competence or low competence
    Moderate competence
    High competence
    Very high competence

16. Accessibility of e-services
    How do you feel about the following statement: the electronic services are not accessible to me e.g. due to my visual impairment
    Completely agree or somewhat agree
    Neither agree nor disagree
    Somewhat disagree or strongly disagree

17. Data security
    How do you feel about the following statement: I am concerned about data security when it comes to my personal details
    Completely agree
    Somewhat agree
    Neither agree nor disagree
    Somewhat disagree
    Strongly disagree

18. Data connections
    How do you feel about the following statement: data connections are poor in my area
    Completely agree or somewhat agree
    Neither agree nor disagree
    Somewhat disagree or strongly disagree

### Diet quality scoring

The diet quality score was constructed from four questions asking the frequency of consumption of roots and vegetables, fish, red meat and fruits and berries. The options for each were: i) never; ii) once a week or more rarely; iii) 2–6 times a week; iv) 1–2 times a day; and v) 3 times a day or more often. The options were graded as shown in table S1, and the final food quality score was obtained by summing the scores from each food category.

| | **i)** | **ii)** | **iii)** | **iv)** | **v)** |
|---|---|---|---|---|---|
| **Vegetables and roots** | 0p | 0p | 1p | 2p | 3p |
| **Fish** | 0p | 1p | 3p | 3p | 3p |
| **Red meat** | 3p | 3p | 2p | 1p | 0p |
| **Fruits and berries** | 0p | 0p | 1p | 2p | 3p |

**Supplementary Table S1.** Scoring of individual food categories
